# Long-Term Mortality and Heart Failure Risk After Pacemaker Implantation: A Nationwide Cohort Study

**DOI:** 10.1101/2025.05.02.25326909

**Authors:** Young Jun Park, Sungjoo Lee, Sungjun Hong, Kyunga Kim, Juwon Kim, Ju Youn Kim, Kyoung-Min Park, Young Keun On, Seung-Jung Park

**Affiliations:** Division of Cardiology, Department of Internal Medicine, Wonju Severance Christian Hospital, Yonsei University Wonju College of Medicine, Wonju, Republic of Korea; Department of Digital Health, Samsung Advanced Institute for Health Sciences & Technology, Sungkyunkwan University, Seoul, Korea; Medical AI Research Center, Research Institute for Future Medicine, Samsung Medical Center, Seoul, Republic of Korea; Biomedical Statistics Center, Research Institute for Future Medicine, Samsung Medical Center, Seoul, Korea; Division of Cardiology, Department of Internal Medicine, Heart Vascular and Stroke Institute, Samsung Medical Center, Sungkyunkwan University School of Medicine, Seoul, Korea

**Keywords:** Pacemaker, Heart failure, Mortality, cardiac resynchronisation therapy, beta-blocker, angiotensin receptor neprilysin inhibitor

## Abstract

**Background:** The long-term risk of heart failure (HF) and mortality following permanent pacemaker (PPM) implantation remains controversial and underexplored.

**Objectives:** This study evaluated these risks and assessed the survival benefits of upgrading to cardiac resynchronisation therapy (CRT) and the impact of standard HF medications.

**Methods:** Using the Korean National Health Insurance Service database, we identified 32,216 patients who underwent PPM implantation without preexisting HF between 2008 and 2019.

**Results:** During the median 3.8-year follow-up period, pacemaker-associated HF (PaHF) and all-cause death occurred in 4170 (12.9%) patients, and 6184 (19.2%) died. PaHF development was strongly associated with all-cause mortality, with a significantly higher risk in the PaHF group than in the non-PaHF group (hazard ratio [HR] 3.11, 95% confidence interval [CI] 2.93–3.32) after adjusting for immortal-time bias and confounders. PaHF incidence and associated mortality were highest within the first 6 months, however, persisted throughout follow-up, with a mortality risk resurgence approximately five years post-implantation. In a propensity score-matched cohort of PaHF patients (n=1,455), CRT-upgrade was associated with a significantly lower risk of mortality (HR 0.34, 95% CI 0.24– 0.47), as were angiotensin receptor-neprilysin inhibitor (ARNI) (HR 0.28, 95% CI 0.14–0.54) and beta-blockers (HR 0.75, 95% CI 0.61–0.93).

**Conclusions:** PaHF development independently predicted mortality post-PPM implantation, while CRT-upgrade and the use of beta-blockers or ARNI were associated with improved survival. Therefore, for PaHF patients, an immediate switching into CRT or conduction system pacing combined with optimal HF medications, may be required to mitigate the ongoing mortality risk.

## Introduction

Permanent pacemaker (PPM) implantation is the standard treatment for patients with symptomatic bradyarrhythmia, including sinus node dysfunction (SND) and atrioventricular block (AVB). While PPM effectively restores cardiac rhythm, it may cause pacing-induced left ventricular (LV) electromechanical dyssynchrony, leading to progressive LV systolic dysfunction and new-onset heart failure (HF), which may increase the risk of mortality.^1–4^ Previous studies have focused primarily on the early incidence of post-PPM HF, affecting approximately 6–25% of patients within 2–4 years. ^1,5–7^ However, the long-term mortality risk associated with chronic ventricular pacing remain insufficiently demonstrated in landmark randomized trials, cohort studies, and meta-analyses.^1,5,6^ ^8–15^ Additionally, many studies are limited by small sample sizes, single-center designs, and short follow-up periods, restricting generalizability and providing an incomplete understanding of long-term outcomes. Moreover, the effectiveness of standard HF medications and the role of cardiac resynchronization therapy (CRT)-upgrades in reducing long-term mortality in patients with post-PPM HF have not been thoroughly evaluated in large, multicenter cohorts.

To address these gaps, we conducted a nationwide cohort study to assess the long-term risk of new-onset HF and its associated mortality following PPM implantation. We further evaluated the impact of standard HF therapies and CRT-upgrades on overall survival in patients with post-PPM HF, aiming to inform optimal management strategies for this patient population.

## Methods

### Data sources

This nationwide cohort study utilized data from the National Health Information Insurance (NHIS) database, a mandatory social health insurance system covering almost the entire South Korean population (over 52 million).^16^ The NHIS database provides comprehensive healthcare information, including demographics, diagnoses (based on the International Classification of Disease, 10th Revision [ICD-10]), hospital visits, admissions, prescription medications (using Anatomical Therapeutic Chemical classification codes), procedures, device usage, and mortality data. The validity and reliability of diagnostic codes in the NHIS have been well-established for major cardiovascular and chronic diseases.^16–20^ This study was approved by the Institutional Review Board of Samsung Medical Center (File-No. 2019-05-075). Informed consent was waived as the data were deidentified in accordance with confidentiality guidelines.

### Study design and population

This study comprised two cohorts (Supplementary Figure1). The primary cohort included patients with de novo PPM implantation and no preexisting HF, allowing evaluation of the incidence of new-onset pacemaker-associated HF (PaHF), and its impact on all-cause mortality. Using procedure and device codes (Supplementary Table 1), we identified 38,921 adult (≥18 years) who underwent PPM implantation between January 2008 and December 2019.

Exclusion criteria were (1) a history of HF before PPM implantation (n=6,389), defined as hospitalization with an HF diagnosis code (I50.9) or prior prescription of angiotensin receptor neprilysin inhibitor (ARNI), (2) reimplantation following PPM system removal (n=892), and (3) death on the day of PPM implantation (n=6). After applying these exclusions, 32,216 patients remained for analysis, of whom 4,170 developed PaHF and 28,046 did not.

The secondary cohort comprised 4,166 patients from the primary cohort who developed PaHF, excluding 4 who died on the day of PaHF diagnosis. This cohort was stratified by treatment strategy into patients with (n=330) and without CRT-upgrade (n=3,836). To emulate a randomized controlled trial, a propensity score (PS)-matched cohort was generated using a 1:4 ratio, resulting in 316 and 1,139 patients in the CRT-upgrade and non-CRT-upgrade groups, respectively.

### Data acquisition

Demographic data, including age and sex, and ICD-10 code-based comorbidities were obtained from claims data collected within one year prior to the index date through to the date of PaHF diagnosis (Supplementary Table 2). Baseline comorbidity burden was represented using the Charlson Comorbidity Index (CCI) (Supplementary Table 3). Medication history was extracted for renin-angiotensin system (RAS) inhibitors, including angiotensin-converting enzyme inhibitors (ACEIs), angiotensin II receptor blockers (ARBs), and ARNI, as well as beta-blockers and mineralocorticoid receptor antagonists (MRAs). Pacemaker type and CRT-upgrade status were identified using ICD-10 codes (Supplementary Table 1).

### Study outcomes and follow-up

The primary outcomes were the incidences of PaHF and all-cause mortality following PPM implantation. PaHF was strictly defined as hospitalization with a newly assigned HF code (I50.9), accompanied by ≥2 HF medication claims (ACEIs/ARBs, beta-blockers, or MRAs). Patients receiving CRT-upgrades or ARNI treatment were also considered as having PaHF. A broad definition of PaHF included all patients with a newly assigned HF code (Supplementary Table 4). All primary analyses used the strict definition.

Censoring for PaHF incidence occurred at death, occurrence of other HF-related diseases not attributable to ventricular pacing (e.g., myocardial infarction, myocarditis, alcoholic cardiomyopathy, cardiac sarcoidosis, or cardiac amyloidosis; Supplementary Table 5), or the last follow-up, whichever occurred first. Follow-up for all-cause mortality was measured from the date of PPM implantation to death or last follow-up. In the secondary PaHF cohort, follow-up for all-cause mortality was measured from the first PaHF diagnosis to death or last follow-up.

### Statistical analyses

Baseline characteristics were summarized as means (standard deviations) or median (interquartile range [IQR]) for continuous variables and counts (percentages) for categorical variables. Continuous variables were compared using the Student’s t-test or Mann–Whitney rank-sum test, while categorical variables were compared using the χ2 test or Fisher exact test, as appropriate. Incidence rates of PaHF and all-cause death were expressed as events per 100 patient-years (PYs) with exact Poisson 95 % confidence intervals (CIs).

The cumulative incidence of PaHF was estimated using the Kaplan-Meier method, while hazard rates (instantaneous incidence rates) were calculated using nonparametric smoothing with B-splines and a generalized linear mixed model across the entire follow-up period.^21^ Multivariable Cox proportional hazards regression was used to estimate hazard ratios (HRs) with 95% CIs for PaHF predictors.

For post-PPM all-cause mortality, the onset of PaHF was treated as a time-dependent covariate in extended Kaplan-Meier estimation and multivariable Cox proportional hazard regression analyses to address immortal-time bias (Supplementary Figure 2).^22,23^ Additionally, multivariable Cox regression with cubic spline functions was employed to examine the time-varying effect of PaHF on all-cause mortality throughout the follow-up period.^24^ Subgroup analyses were conducted, stratified by age, sex, comorbidities, pacemaker type, pacing indication, and medication use, with interaction terms included to evaluate effect modification.

In the PaHF cohort, mortality was compared between patients managed with and without CRT-upgrade, before and after PS-matching. The PS for receiving CRT-upgrade was estimated using multivariable binary logistic regression with potential confounders, and balance between matched groups was evaluated using standardized mean differences (SMDs). Kaplan-Meier survival curves were generated and compared using log-rank and stratified log-rank tests in the entire and the PS-matched PaHF cohorts. Multivariable Cox proportional hazard models were used to evaluate the associations between the CRT-upgrade status and post-PaHF mortality. Covariates for multivariable models were selected based on prior studies and clinical relevance (Supplementary Tables 6). Given the short PaHF-to-CRT-upgrade interval (median 0.0 months, IQR 0.0–2.8), PaHF was not considered a time-varying covariate. Multicollinearity was assessed by variance inflation factor, with values >4 indicating non-negligible collinearity. The proportional hazards assumption was tested using the scaled Schoenfeld residuals. Statistical significance was defined as two-sided P <0.05. All analyses were conducted using SAS version 9.3 (SAS Institute, Cary, NC, USA) and R version 4.1.0 (The R Foundation, www.R-project.org).

## Results

### PaHF and All-cause Mortality Following de novo Pacemaker Implantation

Among the 32,216 PPM patients in the primary cohort, 13,632 (42.3%) were male, 20,246 (63.4%) had AVB, and 27,073 (84.0%) received dual-chamber PPMs. The mean age was 70.6±12.1 years. Detailed baseline characteristics of the study population are presented in Table 1.

**Table 1.** Baseline characteristics of the PPM and PaHF cohorts.

| Variables | PPM Cohort |  |  |  | PaHF Cohort (Propensity Score-Matched) |  |  |
| --- | --- | --- | --- | --- | --- | --- | --- |
|  | Overall<br>(n = 32 216) | PaHF<br>(n = 4170) | No PaHF<br>(n = 28 046) | aSMD | CRT-upgrade*<br>(n = 316) | No-upgrade*<br>(n = 1139) | aSMD |
| *Demographics and medical history* |  |  |  |  |  |  |  |
| Age, years | 70.6 ± 12.1 | 71.9 ± 11.2 | 70.4 ± 12.2 | 0.128 | 67.7 ± 13.4 | 70.3 ± 12.4 | 0.196 |
| Age ≥ 65 years | 24 033 (74.6) | 3314 (79.5) | 20 719 (73.9) | 0.133 | 207 (65.5) | 817 (71.7) | 0.131 |
| Male | 13 632 (42.3) | 1815 (43.5) | 11 817 (42.1) | 0.028 | 149 (47.2) | 524 (46.0) | 0.022 |
| Diabetes | 10 291 (31.9) | 1578 (37.8) | 8713 (31.1) | 0.143 | 84 (26.6) | 342 (30.0) | 0.078 |
| Hypertension | 24 711 (76.7) | 3557 (85.3) | 21 154 (75.4) | 0.250 | 236 (74.7) | 895 (78.6) | 0.089 |
| CAD | 13 003 (40.4) | 2170 (52.0) | 10 833 (38.6) | 0.272 | 174 (55.1) | 651 (57.2) | 0.042 |
| PAD | 7453 (23.1) | 1015 (24.3) | 6438 (23.0) | 0.033 | 111 (35.1) | 418 (36.7) | 0.033 |
| CKD/ESRD | 2263 (7.0) | 390 (9.4) | 1873 (6.7) | 0.099 | 39 (12.3) | 168 (14.7) | 0.073 |
| VHD | 2976 (9.2) | 631 (15.1) | 2345 (8.4) | 0.211 | 54 (17.1) | 176 (15.5) | 0.043 |
| AF | 7297 (22.7) | 1169 (28.0) | 6128 (21.8) | 0.143 | 116 (36.7) | 439 (38.5) | 0.038 |
| COPD | 6294 (19.5) | 1007 (24.1) | 5287 (18.9) | 0.129 | 111 (35.1) | 419 (36.8) | 0.035 |
| CCI ≥ 3 unit | 19 612 (60.9) | 2885 (69.2) | 16 727 (59.6) | 0.200 | 260 (82.3) | 994 (87.3) | 0.131 |
| *Pacemaker-related variables* |  |  |  |  |  |  |  |
| AV block | 20 246 (63.4) | 2502 (60.0) | 17 744 (63.3) | 0.068 | 226 (71.5) | 790 (69.4) | 0.048 |
| SND | 11 675 (36.6) | 1631 (39.1) | 10 044 (35.8) |  | 90 (28.5) | 349 (30.6) |  |
| Dual-chamber | 27 073 (84.0) | 3136 (75.2) | 23 937 (85.3) | 0.262 | 276 (87.3) | 987 (86.7) | 0.021 |
| ***Medications*** |  |  |  |  |  |  |  |
| ACEIs or ARBs | 21 490 (66.7) | 3309 (79.4) | 18 181 (64.8) | 0.328 | 209 (66.1) | 786 (69.0) | 0.061 |
| ARNI | - | - | - | - | 19 (6.0) | 126 (11.1) | 0.212 |
| Beta blockers | 13 905 (43.2) | 2390 (57.3) | 11 515 (41.1) | 0.330 | 141 (44.6) | 523 (45.9) | 0.026 |
| MRAs | 5274 (16.4) | 1222 (29.3) | 4072 (14.5) | 0.363 | 97 (30.7) | 359 (31.5) | 0.018 |
| Loop diuretics | 12 611 (39.1) | 2389 (57.3) | 10 222 (36.4) | 0.427 | 135 (42.7) | 610 (53.6) | 0.219 |

Values are expressed as means ± standard deviations or n (%).
Abbreviations: PPM, permanent pacemaker; PaHF, pacemaker-associated heart failure; aSMD, absolute standardised mean difference; CRT, cardiac resynchronization therapy; CAD, coronary artery disease; PAD, peripheral artery disease; CKD, chronic kidney disease; ESRD, end-stage renal disease; VHD, valvular heart disease; AF, atrial fibrillation; COPD, chronic obstructive pulmonary disease; CCI, Charlson comorbidity index; AV block, atrioventricular block; SND, sinus node dysfunction; ACEIs, angiotensin-converting enzyme inhibitors; ARBs, angiotensin II receptor antagonists; MRAs, mineralocorticoid receptor antagonists.
\*, The CRT-upgrade group included patients who received CRT-upgrade in addition to HF medications, while the No-upgrade group comprised those managed with HF medications alone.

Over a median follow-up of 3.8 years (IQR, 1.7−6.7), PaHF occurred in 4,170 patients (12.9%) and all-cause mortality in 6,184 patients (19.2%) of 32,216 PPM recipients, with incidence rates of 3.3 (95% CI, 3.2−3.4) and 4.4 (95% CI, 4.3−4.5) per 100 PYs, respectively (Supplementary Figure 1). The median time to PaHF and death was 2.2 years (IQR, 0.7−4.8) and 3.0 years (IQR, 1.2−5.4) following PPM implantation, respectively.

Patients who developed PaHF had worse baseline characteristics, including older age, higher comorbidity burden, and greater medication use (Table 1). These patients also exhibited significantly higher all-cause mortality compared to those without PaHF: 6.2 [95% CI, 5.9−6.5] vs. 4.0 [95% CI, 3.9−4.1] per 100 PYs, P<0.001). Extended Kaplan-Meier curves also indicated significantly worse prognosis in the PaHF group (Figure 1A). Subgroup analyses demonstrated a consistently higher mortality risk across all strata for PaHF patients (Figure 1B). Multivariable Cox proportional hazard regression identified PaHF development as an independent predictor for all-cause mortality post-PPM (HR 3.11, 95% CI, 2.93−3.32, P<0.001), even after adjusting for immortal-time bias and other potential confounders (Supplementary Table 7).

**Figure 1.**
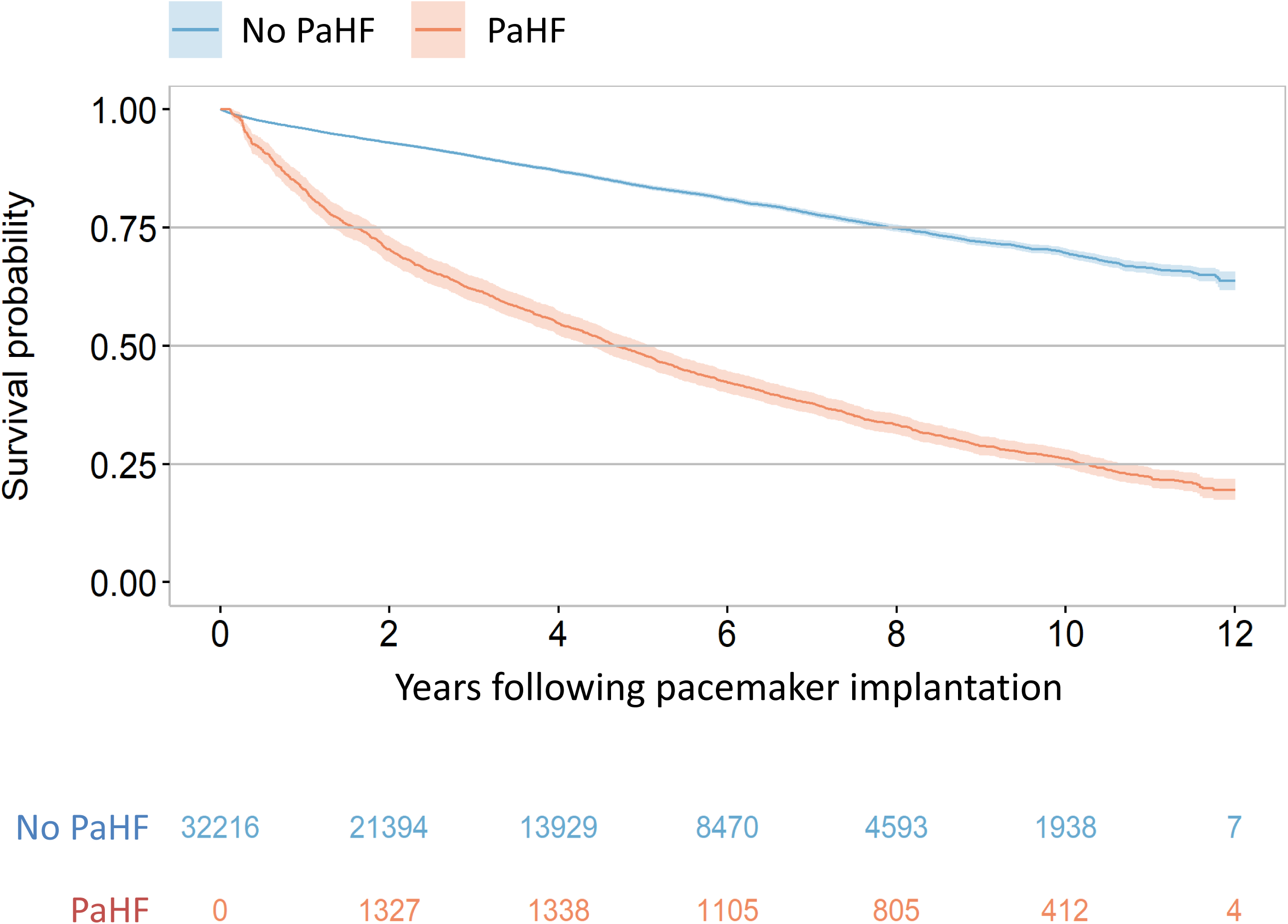
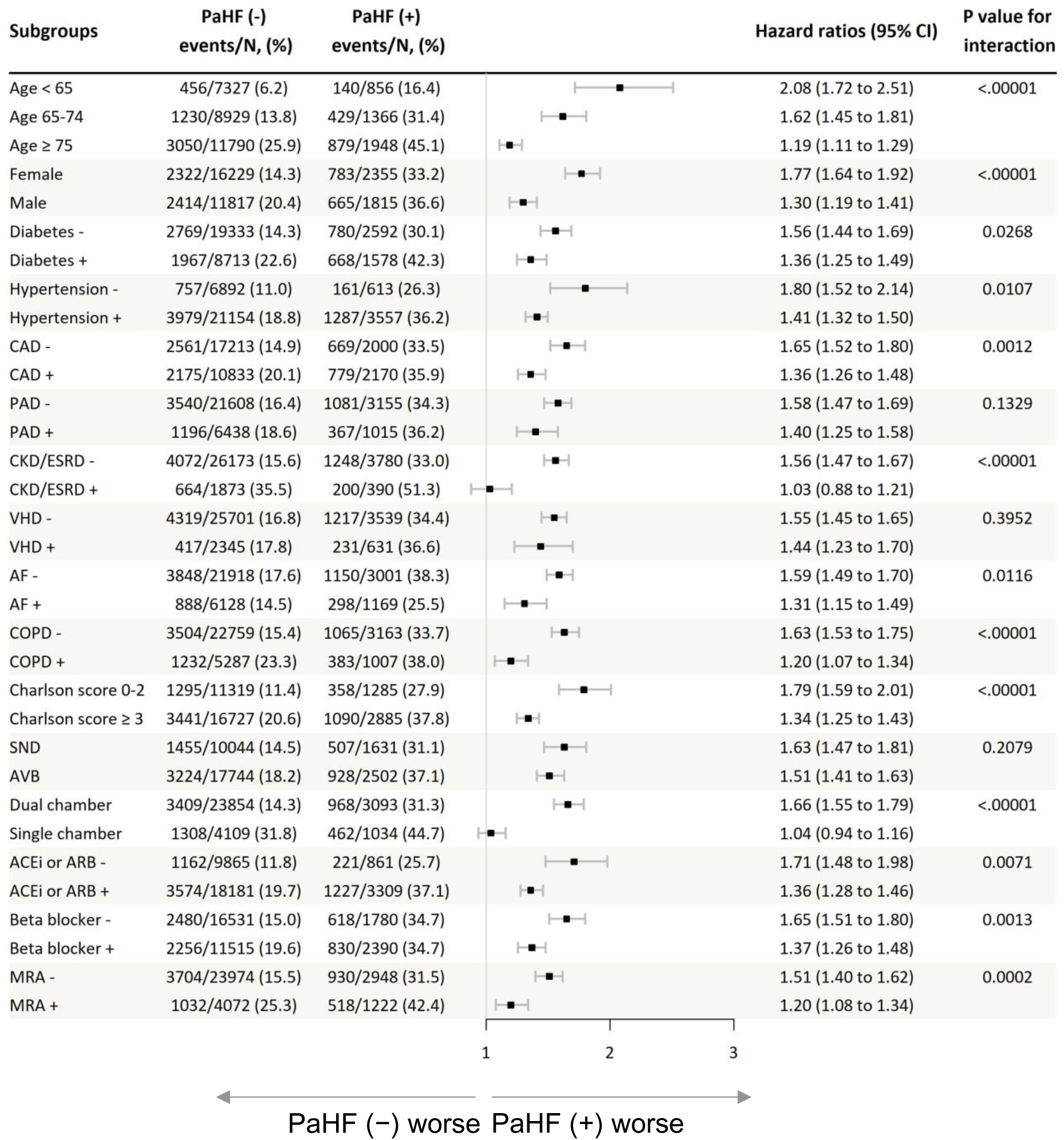
Kaplan–Meier Survival Curves and Subgroup Analysis. (A) Extended Kaplan–Meier survival curves for all-cause mortality show that patients who developed PaHF exhibited significantly worse survival outcomes than those who did not. The curves were adjusted for the time from pacemaker implantation to PaHF diagnosis, treated as a time-dependent covariate. (B) The forest plot of the subgroup analysis demonstrates that across all subgroups, patients with PaHF consistently had a higher risk of mortality compared to those without. ***Abbreviations***: PaHF, pacemaker-associated heart failure; CI, confidence interval; CAD, coronary artery disease; PAD, peripheral artery disease; CKD, chronic kidney disease; ESRD, end-stage renal disease; VHD, valvular heart disease; AF, atrial fibrillation; COPD, chronic obstructive pulmonary disease; SND, sinus node dysfunction; AVB, atrioventricular block; ACEI, angiotensin-converting enzyme inhibitor; ARB, angiotensin receptor blocker; MRA, mineralocorticoid receptor antagonist.

### Temporal Trends in PaHF Risk and Associated Mortality

The hazard rates of strictly and broadly defined PaHFs, representing instantaneous incidence at specific time points, were highest within the first 6 months following PPM implantation, but remained above zero throughout the follow-up period, forming L-shaped curves (Figure 2A). The time-varying mortality risk associated with PaHF, estimated using multivariable Cox regression with cubic splines, also peaked within the first 6 months, declined to its nadir approximately 5 years post-implantation, and subsequently increased again over time, forming a U-shaped curve (Figure 2B).

**Figure 2.**
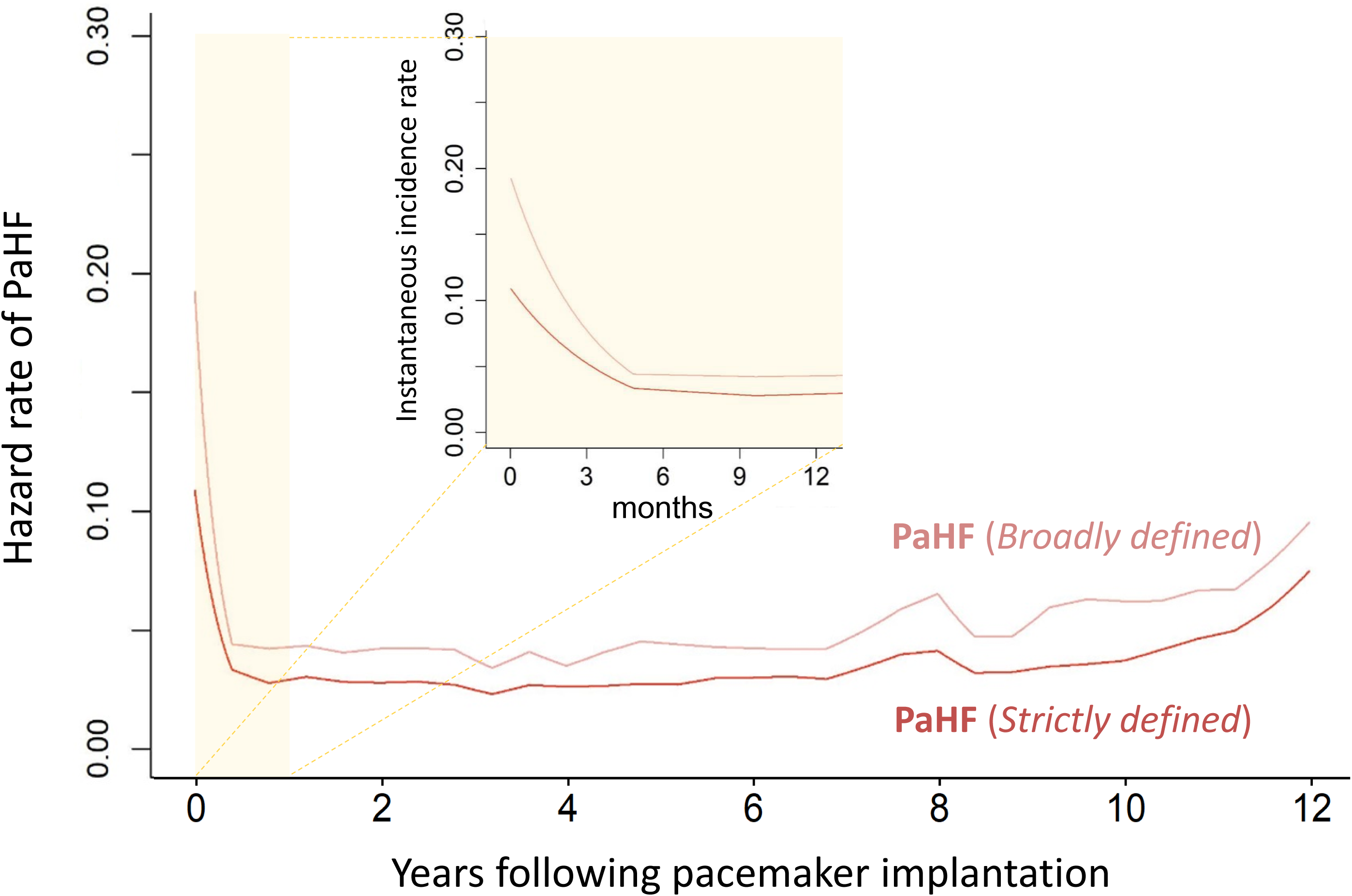
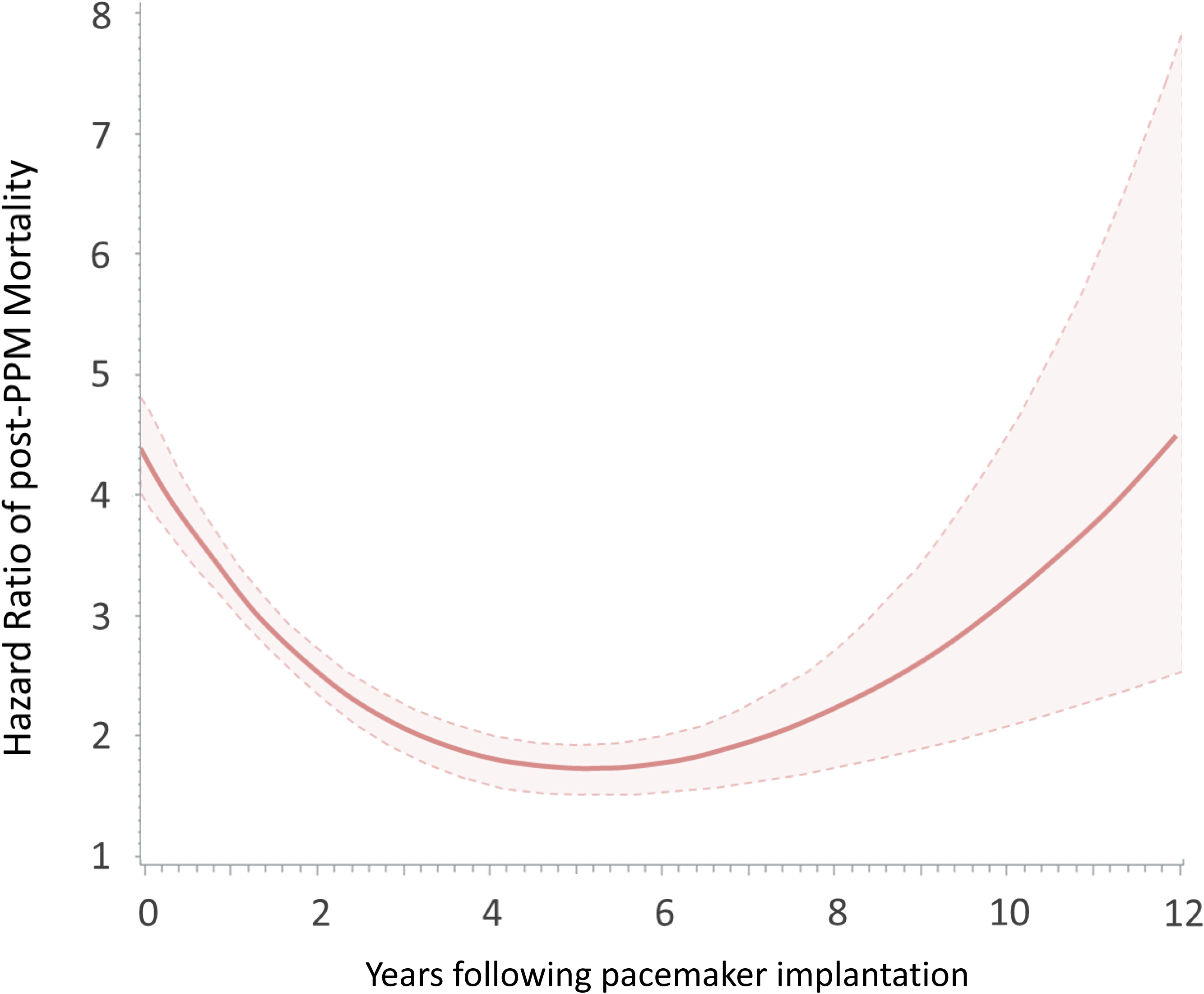
Hazard Rates of PaHF and Time-Varying Mortality Risk. (A) The hazard rates of strictly and broadly defined PaHFs, representing instantaneous incidence at specific time points, peaked within the first 6 months after PPM implantation and remained elevated throughout the follow-up period. The inset illustrates the rapid decline in hazard rates within the first year. (B) The time-varying mortality risk post-PPM implantation, estimated using multivariable Cox regression with cubic splines, was highest within the first 6 months, reached its lowest point around 5 years, and rose again thereafter. The shaded regions indicate 95% confidence intervals, reflecting the variability in HR estimates over time. ***Abbreviations***: PaHF, pacemaker-associated heart failure; PPM, permanent pacemaker.

### Mortality Determinants in PaHF: CRT-Upgrade, HF Medications, and Clinical Predictors

Baseline characteristics of the PS-matched PaHF cohort at the time of PaHF diagnosis were well-balanced between the CRT-upgrade and non-upgrade groups, except for age ≥65 years and use of ARNI or loop diuretics (Table 1). However, all-cause mortality rates were significantly lower in the CRT-upgrade group than in the non-upgrade group (4.1 [95% CI 3.0−5.5] vs. 11.4 [95% CI 10.3−12.6] per 100 PYs, P<0.001). Kaplan-Meier analyses confirmed better survival for PaHF patients with CRT-upgrade compared to those without, both before and after PS-matching (Figures 3A and 3B).

**Figure 3.**
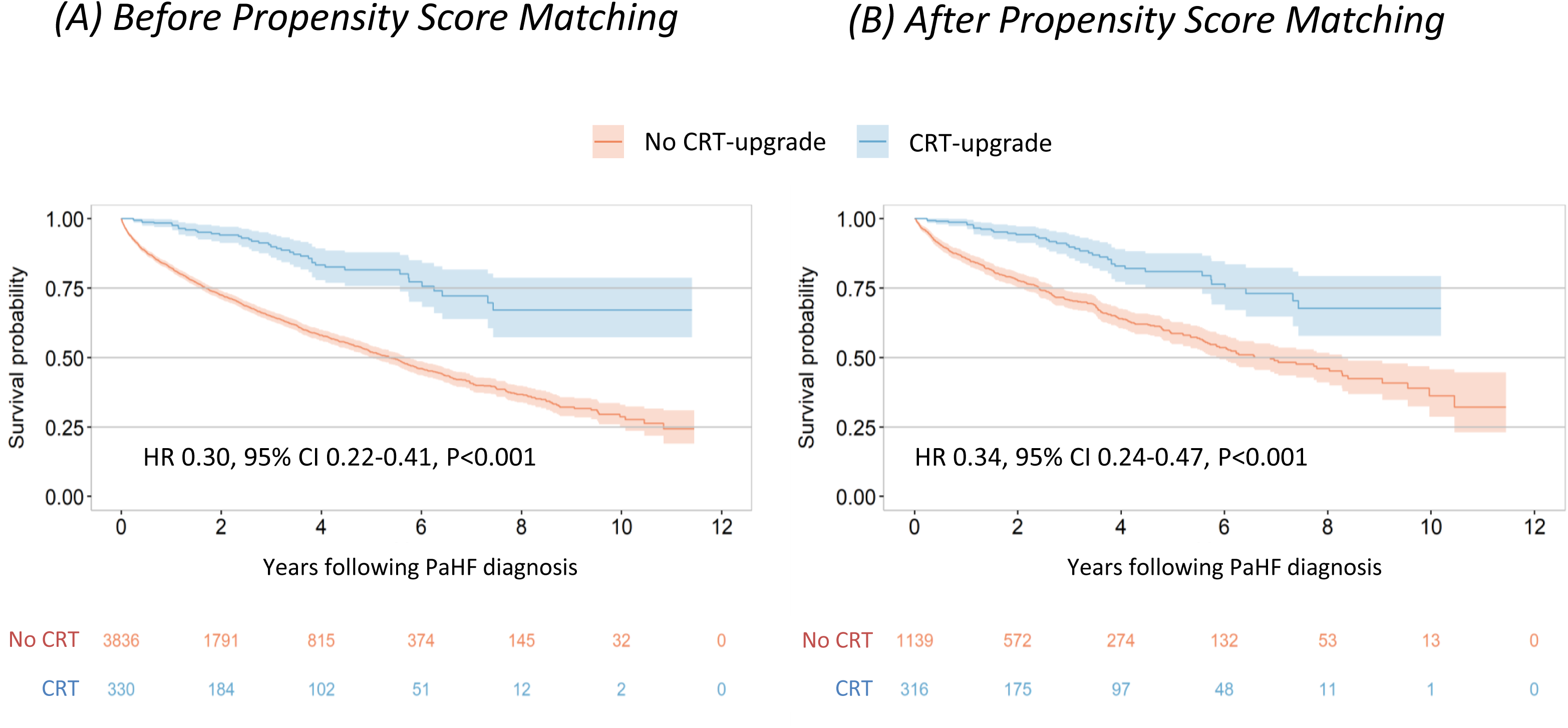
Kaplan–Meier Survival Curves for PaHF Patients With and Without CRT-Upgrade. Long-term prognosis was significantly improved for patients who underwent CRT-upgrade in addition to medical therapy compared to those receiving medical therapy alone, without CRT-upgrade, in both the entire cohort (A) and the propensity score-matched cohort (B). ***Abbreviations***: CRT, cardiac resynchronization therapy; PaHF, pacemaker-associated heart failure. **Central illustration**. Clinical Outcomes and Mortality Risk Factors Following Permanent Pacemaker Implantation The study analyzed 32,216 patients who received permanent pacemaker (PPM) implantation without preexisting heart failure, of whom 4,170 developed pacemaker-associated heart failure (PaHF) while 28,046 did not during a median follow-up of 3.8 years. The survival analysis showed significantly worse outcomes in the PaHF group compared to the non-PaHF group. Risk factors for mortality in PaHF patients included age ≥65 years, male sex, diabetes, and CKD/ESRD. In contrast, protective factors against mortality included CRT-upgrade and optimized heart failure medications, particularly ARNI (HR 0.28) and β-blockers (HR 0.75). In the propensity-matched PaHF population, CRT-upgrade showed significantly better survival compared to medical treatment alone. ***Abbreviations*:** CKD, chronic kidney disease; CRT, cardiac resynchronization therapy; ESRD, end-stage renal disease; HF, heart failure; PaHF, pacemaker-associated heart failure; PPM, pacemaker.

Among patient factors, advanced age, male, DM, and CKD/ESRD were significantly associated with increased all-cause mortality. In contrast, CRT-upgrade, RAS inhibitors, and beta-blockers were independent protective factors, with CRT-upgrade showing the strongest protective effect (HR 0.34, 95% CI 0.24−0.47, P<0.001; model 1, Table 2). However, when RAS inhibitors were categorized as ACEIs/ARBs versus ARNI, only ARNI demonstrated a significant protective effect (HR 0.28, 95% CI 0.14−0.54, P <0.001; model 2).

**Table 2.**
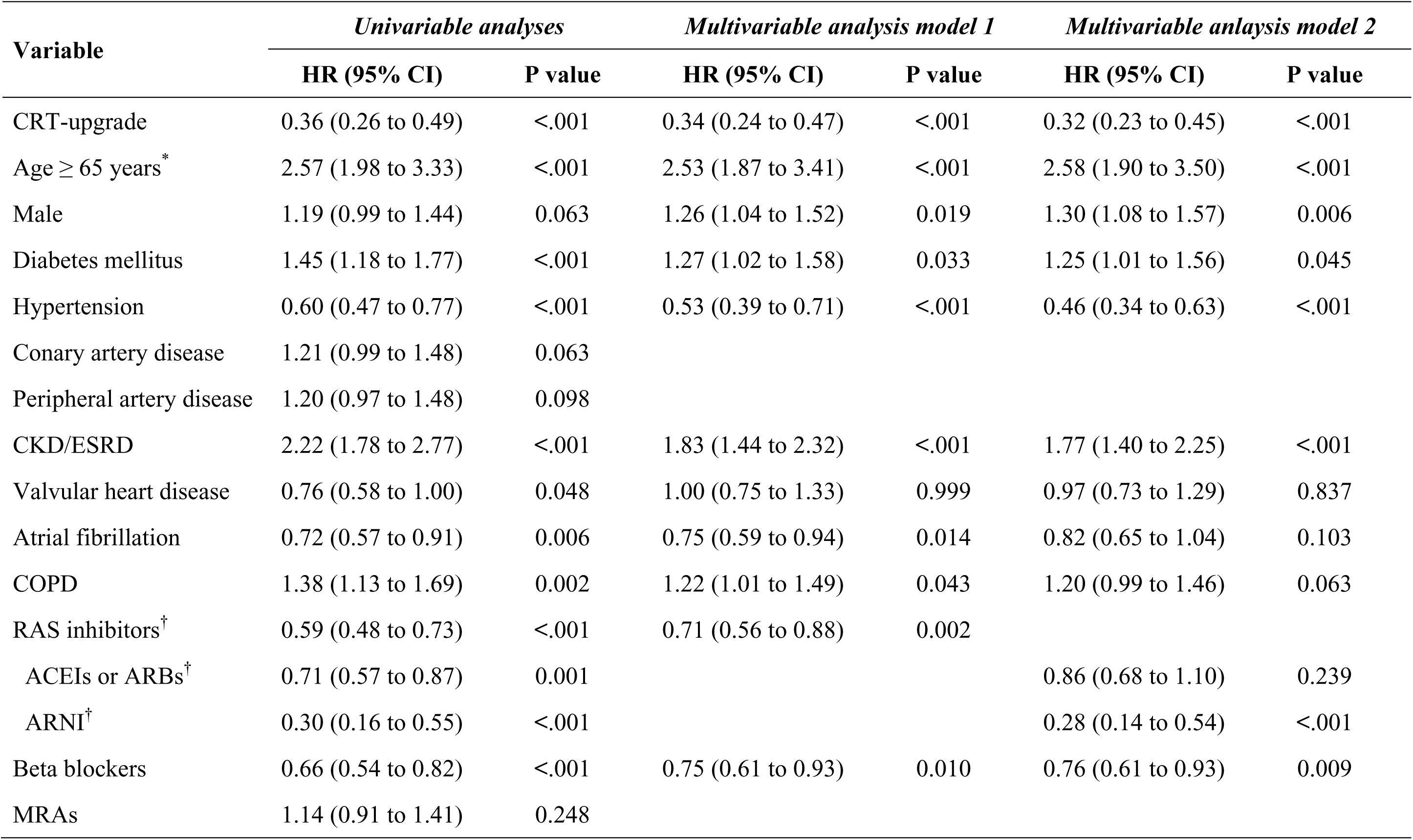
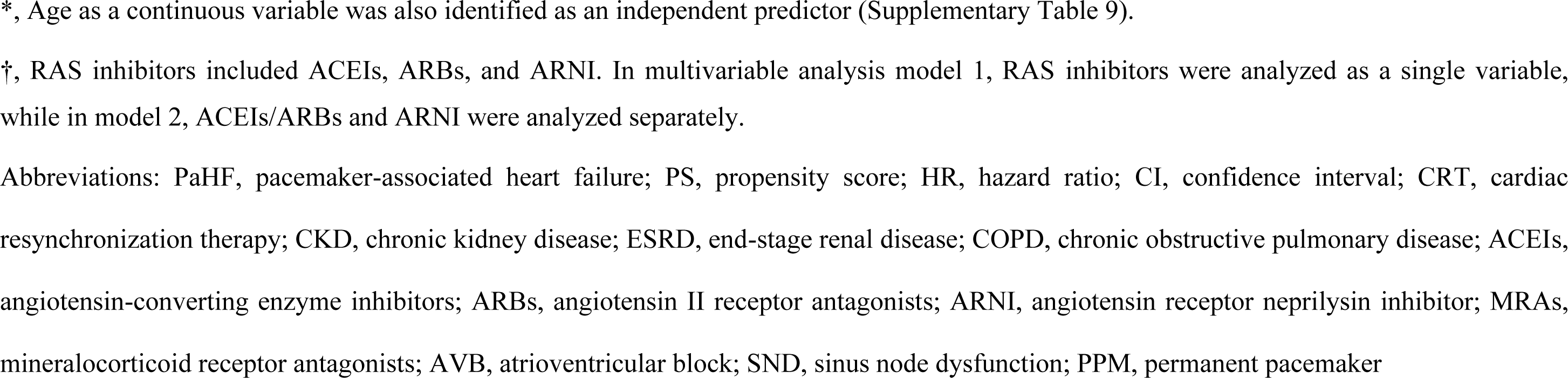
Independent risk factors of all-cause mortality in the propensity score-matched PaHF cohort.

| Variable | *Univariable analyses* |  | *Multivariable analysis model 1* |  | *Multivariable analysis model 2* |  |
| --- | --- | --- | --- | --- | --- | --- |
|  | HR (95% CI) | P value | HR (95% CI) | P value | HR (95% CI) | P value |
| CRT-upgrade | 0.36 (0.26 to 0.49) | <.001 | 0.34 (0.24 to 0.47) | <.001 | 0.32 (0.23 to 0.45) | <.001 |
| Age ≥ 65 years* | 2.57 (1.98 to 3.33) | <.001 | 2.53 (1.87 to 3.41) | <.001 | 2.58 (1.90 to 3.50) | <.001 |
| Male | 1.19 (0.99 to 1.44) | 0.063 | 1.26 (1.04 to 1.52) | 0.019 | 1.30 (1.08 to 1.57) | 0.006 |
| Diabetes mellitus | 1.45 (1.18 to 1.77) | <.001 | 1.27 (1.02 to 1.58) | 0.033 | 1.25 (1.01 to 1.56) | 0.045 |
| Hypertension | 0.60 (0.47 to 0.77) | <.001 | 0.53 (0.39 to 0.71) | <.001 | 0.46 (0.34 to 0.63) | <.001 |
| Coronary artery disease | 1.21 (0.99 to 1.48) | 0.063 |  |  |  |  |
| Peripheral artery disease | 1.20 (0.97 to 1.48) | 0.098 |  |  |  |  |
| CKD/ESRD | 2.22 (1.78 to 2.77) | <.001 | 1.83 (1.44 to 2.32) | <.001 | 1.77 (1.40 to 2.25) | <.001 |
| Valvular heart disease | 0.76 (0.58 to 1.00) | 0.048 | 1.00 (0.75 to 1.33) | 0.999 | 0.97 (0.73 to 1.29) | 0.837 |
| Atrial fibrillation | 0.72 (0.57 to 0.91) | 0.006 | 0.75 (0.59 to 0.94) | 0.014 | 0.82 (0.65 to 1.04) | 0.103 |
| COPD | 1.38 (1.13 to 1.69) | 0.002 | 1.22 (1.01 to 1.49) | 0.043 | 1.20 (0.99 to 1.46) | 0.063 |
| RAS inhibitors<sup>†</sup> | 0.59 (0.48 to 0.73) | <.001 | 0.71 (0.56 to 0.88) | 0.002 |  |  |
| ACEIs or ARBs<sup>†</sup> | 0.71 (0.57 to 0.87) | 0.001 |  |  | 0.86 (0.68 to 1.10) | 0.239 |
| ARNI<sup>†</sup> | 0.30 (0.16 to 0.55) | <.001 |  |  | 0.28 (0.14 to 0.54) | <.001 |
| Beta blockers | 0.66 (0.54 to 0.82) | <.001 | 0.75 (0.61 to 0.93) | 0.010 | 0.76 (0.61 to 0.93) | 0.009 |
| MRAs | 1.14 (0.91 to 1.41) | 0.248 |  |  |  |  |

\*, Age as a continuous variable was also identified as an independent predictor (Supplementary Table 9).
†, RAS inhibitors included ACEIs, ARBs, and ARNI. In multivariable analysis model 1, RAS inhibitors were analyzed as a single variable, while in model 2, ACEIs/ARBs and ARNI were analyzed separately.
Abbreviations: PaHF, pacemaker-associated heart failure; PS, propensity score; HR, hazard ratio; CI, confidence interval; CRT, cardiac resynchronization therapy; CKD, chronic kidney disease; ESRD, end-stage renal disease; COPD, chronic obstructive pulmonary disease; ACEIs, angiotensin-converting enzyme inhibitors; ARBs, angiotensin II receptor antagonists; ARNI, angiotensin receptor neprilysin inhibitor; MRAs, mineralocorticoid receptor antagonists; AVB, atrioventricular block; SND, sinus node dysfunction; PPM, permanent pacemaker

### Sensitivity analyses

Using the broad PaHF definition, 6,118 patients (19.0%) were identified to develop PaHF, yielding a higher incidence rate of 4.9 per 100 PYs (95% CI 4.8−5.0) compared to the strict definition (Supplementary Figure 3). Sensitivity analyses based on the broad PaHF definition yielded results consistent with the primary findings, demonstrating significantly higher mortality in the PaHF group compared to the non-PaHF group (Supplementary Figures 4 and 5). Subgroup analyses further confirmed these results across all strata (Supplementary Figure 6).

## Discussion

### Main Findings

The primary findings of this study are: (1) PaHF occurred in 12.9% and 19.0% of PPM patients based on the strict and broad definitions, respectively, over a median follow-up of 3.8 years; (2) the overall post-PPM mortality rate was 19.2%, with PaHF development identified as an independent predictor for all-cause mortality, carrying an approximately threefold increased risk compared to those without PaHF after adjusting for immortal-time bias and potential confounders; (3) the PaHF incidence and associated mortality rates were highest within the first 6 months, however, persisted throughout the follow-up period, with a resurgence in mortality risk approximately five years post-implantation; and (4) CRT-upgrade, RAS inhibitors, and beta-blockers were associated with improved overall survival in PaHF patients, with ARNI more likely effective than ACEIs/ARBs in reducing mortality.

### PaHF Incidence and Persistent Mortality Risk Following PPM Implantation

The incidence of PaHF in our study aligns with previous reports. A large single-center study reported a 12.3% incidence of PaHF based on the occurrence of moderate to severe LV systolic dysfunction, in line with our strict PaHF definition.^2^ Conversely, the nationwide MarketScan study, using a broader definition based solely on HF codes, estimated a higher incidence of 25.8%.^25^

Despite numerous studies linking chronic ventricular pacing to an elevated HF risk, its long-term impact on mortality remains inconclusive, particularly in patients without preexisting HF. The randomized MOST trial demonstrated that a higher ventricular pacing burden (>40%) markedly increased the risk of HF hospitalization, but did not assess mortality outcomes.^26^ Similarly, a recent Danish registry study primarily focused on the incidence and predictors of PaHF, leaving mortality risk underexplored.^27^ Even landmark trials like the randomized DAVID, SAVE PACe, DANPACE, and studies failed to demonstrate a significant association between pacing burden and mortality (P=0.15 and P=0.53, respectively).^8–10^ A German registry study likewise found no difference in mortality between patients with AVB and those with SND (17% vs. 17%).^5^ Additionally, chronic ventricular pacing did not emerge as an independent predictor of reduced survival in a study of patients with isolated congenital AVB.^11^ Furthermore, pacing-minimization algorithms have consistently failed to reduce all-cause mortality in randomized studies and meta-analyses.^9,12–15^ A likely explanation for these inconclusive association between chronic ventricular pacing and mortality risk may be the relatively short follow-up periods in these studies, which averaged 2.5 years or less.^5,8,9,12–15^ Such durations may be insufficient to detect the long-term mortality effects of chronic pacing or pacing-minimization algorithms. In contrast, our study offers a more extended follow-up of 4.4±3.2 years, with mean intervals of 3.0±2.8 years from PPM implantation to PaHF onset and 3.6±2.8 years to death, clearly demonstrating that PaHF development increased mortality risk following PPM implantation. Notably, our data revealed that PaHF-associated mortality risk remained elevated throughout the follow-up, forming a U-shaped curve with an early peak, a nadir around five years, and a resurgence thereafter, suggesting that pacing-related mortality risk persists unabated over time, a pattern not captured in studies with shorter follow-up periods (Figure 1C)

### Clinical Predictors of Mortality in PaHF Patients

Consistent with prior reports, CKD/ESRD, male, and advanced age were associated with increased mortality in PaHF patients (Table 2). In CKD, conditions such as ventricular hypertrophy, myocardial fibrosis, and uremia can impair myocardial conduction velocity^28,29^, increasing pacing-induced dyssynchrony and mortality risk.^6,27,30^ The higher mortality observed in men may be attributable to their larger myocardial mass, which requires more time for single-site stimulation to activate the entire heart, leading to more severe dyssynchrony.^31,32^ This greater susceptibility to PaHF in men with larger hearts may reflect a counterpart phenomenon to the superior CRT response seen in women with smaller hearts.^33^ The specific role of DM on PaHF occurrence or mortality has rarely been explored. However, DM is frequently associated with advanced age, CKD/ESRD, and myocardial infarction, potentially compounding mortality risk.^34,35^

### Prevention, Monitoring, and Management of PaHF and Its Mortality Risk

When PPM implantation is indicated for patients with preserved EF or no preexisting HF, prophylactic CRT implantation may not be justified, given the relatively low PaHF incidence observed in our data (12.9%) and in recent large registry-based studies (10.6% and 25.8%).^25,27^ Instead, cardiac conduction system pacing may offer a better alternative to conventional PPM or prophylactic CRT for patients with multiple risk factors of PaHF, particularly for younger patients with a high predicted pacing burden and longer expected lifespan.^36,37^ Indeed, our data suggest that the relative contribution of PaHF to mortality is increasingly accentuated in younger age groups (P_interaction_<0.001, Figure 1B).

After PPM implantation, there are currently no established guidelines for the timing or frequency of PaHF monitoring. Tayal et al. suggested that echocardiographic evaluations within 6 months post-PPM could identify the majority of PaHF cases, based on their observation of peak incidence within this timeframe.^27^ Our data corroborate these findings, as both PaHF incidence and its related mortality were highest within the first 6 months. However, a resurgence of mortality risk was observed approximately 5 years post-implantation. Furthermore, risk factors such as advanced age, DM, CKD/ESRD, and atrial fibrillation may accumulate over time, further elevating the risk of PaHF and mortality. Therefore, regular cardiac assessments may be warranted, even beyond this initial period.

Once PaHF is diagnosed, it remains unclear whether CRT-upgrade should be performed immediately or after a period of medical therapy. A recent report from Duke University reported that patients with left bundle branch block and reduced EF, showed no improvement or even worsened LV function despite 3-to-6 months of guideline-directed medical therapy, raising question about the appropriateness of routine pre-CRT HF medications in the presence of left bundle branch block.^38^ Likewise, the benefit of pre-CRT medical therapy was not evident in pacing-dependent HF patients with similar dyssynchrony.^7,39^ Our study also showed that MRAs and ACEIs/ARBs were not significantly associated with reduced mortality in PaHF patients, while ARNI and beta-blockers provided mortality benefit (Table 2). These results support recent guidelines recommending ARNI over ACEIs/ARBs for HF with reduced EF.^40,41^ Furthermore, CRT-upgrade combined with medical therapy significantly improved survival compared to medical therapy alone, with Kaplan-Meier survival curves diverging early after PaHF diagnosis (Figure 3). Accordingly, immediate CRT-upgrade or switching to conduction system pacing, along with optimized HF medications, preferably ARNI and beta-blockers, may be a more reasonable strategy than delayed upgrade.

### Limitations

We acknowledge several limitations. First, echocardiographic and device interrogation data were lacking for this cohort, limiting our ability to directly attribute PaHF to RV pacing-induced dyssynchrony. However, in Korea, CRT-upgrades are reimbursed only when the pacing burden exceeds 40%, suggesting that our PaHF patients likely had significant pacing exposure. Additionally, the improved survival observed in PaHF patients following CRT-upgrade further supports the notion that chronic ventricular pacing adversely affected these cases. Second, the superiority of ARNIs over ACEIs/ARBs in reducing PaHF mortality remains preliminary, as randomized controlled trials are needed to validate these findings. Finally, the effect of sodium-glucose cotransporter-2 inhibitors on mortality was not evaluated due to the limited number of PaHF patients receiving these agents.

Despite these limitations, our study may offer valuable insights into the long-term mortality risk following PPM implantation across the entire follow-up period, the optimal timing for CRT-upgrade or switching to conduction system pacing, and the selection of more suitable HF medications for patients with PaHF.

### Conclusions

Our nationwide cohort study found that PaHF development was closely associated with increased mortality following PPM implantation while CRT-upgrade, beta-blockers, and ARNI were strong protective factors for all-cause mortality in PaHF patients. The PaHF incidence and its mortality risk were highest during the early post-PPM period, but likely to persist and resurge over time. These findings underscore the need for regular cardiac function assessments post-PPM implantation. Furthermore, once PaHF is detected, timely transition to cardiac physiological pacing modalities should be considered, along with optimized HF medications.

## Data Availability

This is a study using Korean NHI data, and analysis is possible with government permission.

## Acknowledgement

We thank Mi Yang (Seoul Mental Health Welfare Center) for her great help in retrieving the raw data, and Professor Minsu Park (Department of Information and Statistics, Chungnam National University) for his expert opinions on the study design in the early stage of this study.

## Funding

None declared.

## Disclosure of interest

S.J.-P. received research grants from Boston Scientific, Biotronik, Abbott, and Medtronic. K.-P. received research grants from Boston Scientific. Y.K.-O. received research grants from Bayer AG, Daiichi Sankyo Company. All other authors declare no conflicts of interest.

---

**Central illustration.**
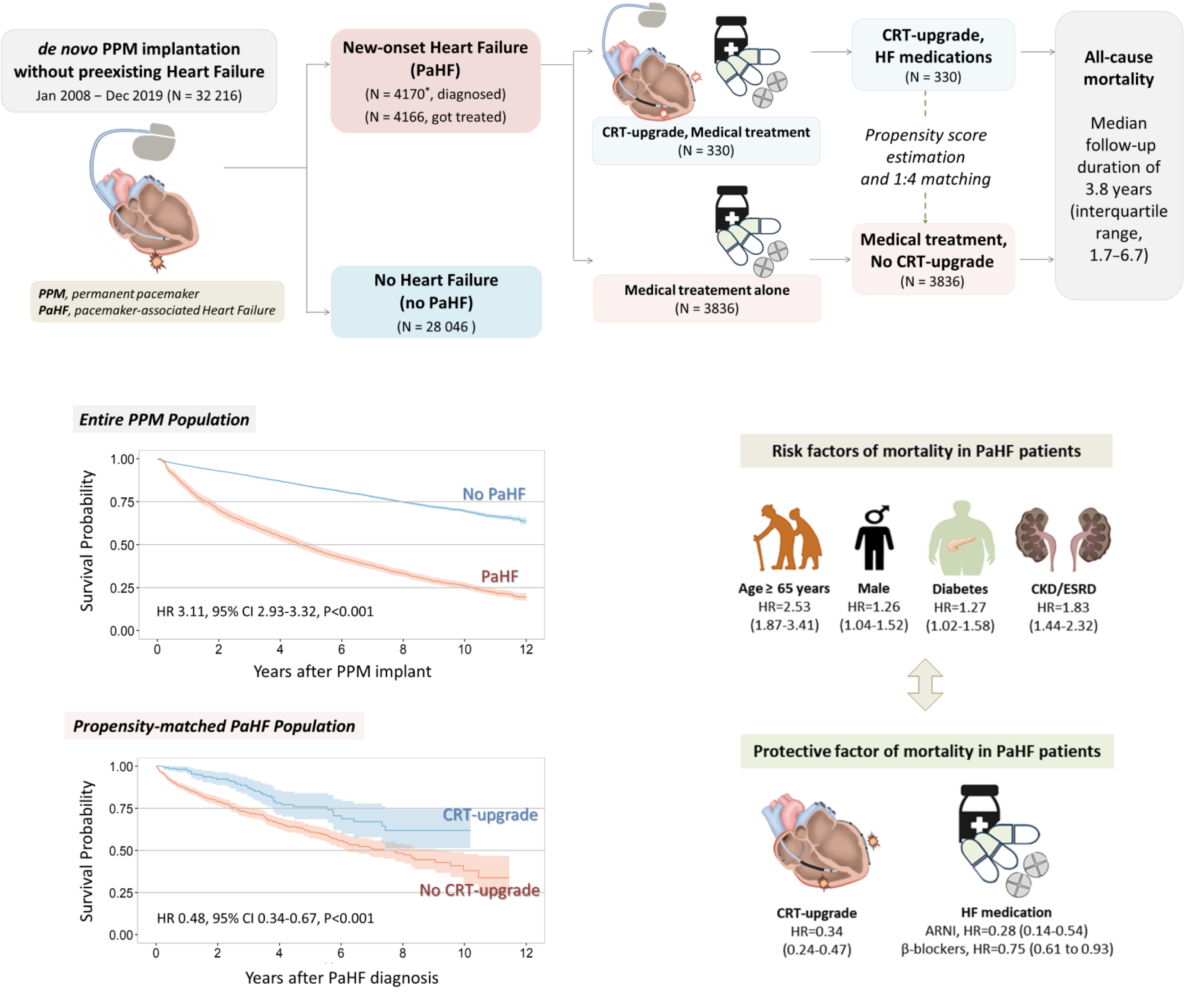

**Supplementary Figure 1. Study profile**

In the primary PPM cohort, 32,216 patients remained for analysis, of whom 4,170 developed PaHF and 28,046 did not. The secondary PaHF cohort was stratified by treatment strategy into patients with (n=330) and without CRT-upgrade (n=3,836). From this, a propensity score-matched cohort was generated using a 1:4 ratio by greedy matching without replacement, applying a caliper width equal to 0.25 times the pooled standard deviation of the logit of the propensity score, resulting in 316 and 1,139 patients in the CRT-upgrade and non-CRT-upgrade groups, respectively.

***Abbreviations*:** PPM, permanent pacemaker; HF, heart failure; PaHF, pacemaker-associated heart failure; CRT, cardiac resynchronization therapy. *, 4 patients died on the day of PaHF diagnosis.

**Supplementary Figure 2. The handling of immortal-time bias in the primary PPM cohort.**

Immortal time refers to the duration during which the outcome could not have occurred prior to PaHF diagnosis. In this analysis, PaHF was treated as a time-dependent covariate in the extended Kaplan-Meier estimator and multivariable Cox proportional hazard regression analysis to mitigate this bias. The extended Kaplan-Meier estimator updates the cohort classification at each time of PaHF development, allowing the PaHF group to contribute to the non-PaHF group until the point of PaHF diagnosis.

***Abbreviations*:** PaHF, pacemaker-associated heart failure; PPM, permanent pacemaker.

**Supplementary Figure 3. Cumulative incidence rates of PaHF development according to strict and broad definitions.**

***Abbreviations*:** PaHF, pacemaker-associated heart failure.

**Supplementary Figure 4. Extended Kaplan–Meier survival curves for all-cause mortality in PPM cohort according to the occurrence of broadly-defined PaHF. *Abbreviations*:** PaHF, pacemaker-associated heart failure.

**Supplementary Figure 5. Time-varying mortality risk of broadly-defined PaHF over the entire follow-up period.**

Hazard ratios across time were estimated using a multivariable Cox regression analysis with a cubic spline function. Solid and dashed lines refer to hazard ratio and its 95% confidence interval, respectively.

***Abbreviations*:** PaHF, pacemaker-associated heart failure; PPM, permanent pacemaker.

**Supplementary Figure 6. Subgroup analysis for all-cause mortality and patients with and without broadly-defined PaHF**

***Abbreviations*:** PaHF, pacemaker-associated heart failure; CI, confidence interval; CAD, coronary artery disease; PAD, peripheral artery disease; CKD, chronic kidney disease; ESRD, end-stage renal disease; VHD, valvular heart disease; AF, atrial fibrillation; COPD, chronic obstructive pulmonary disease; SND, sinus node dysfunction; AVB, atrioventricular block; ACEI, angiotensin-converting enzyme inhibitor; ARB, angiotensin receptor blocker; MRA, mineralocorticoid receptor antagonist.

## Abbreviations

ACEi: Angiotensin-converting-enzyme inhibitors
ARB: Angiotensin II receptor antagonists
ARNI: Angiotensin receptor neprilysin inhibitor
AVB: Atrioventricular block
CRT: Cardiac resynchronization therapy
EF: Ejection fraction
LV: Left ventricular
MRAs: Mineralocorticoid receptor antagonists
PaHF: Pacemaker-associated heart failure
PPM: Permanent pacemaker
SND: Sinus node dysfunction

## Notes

### Clinical Trial

This cohort study was approved by the institutional IRB and Korea NHI.

### Funding Statement

No funding

### Author Declarations

This study was approved by the Institutional Review Board of Samsung Medical Center (File-No. 2019-05-075).

## References

1. Cho SW, Gwag HB, Hwang JK, Chun KJ, Park KM, On YK, Kim JS, Park SJ. Clinical features, predictors, and long-term prognosis of pacing-induced cardiomyopathy. Eur J Heart Fail 2019;21:643–651. doi: 10.1002/ejhf.1427

2. Kiehl EL, Makki T, Kumar R, Gumber D, Kwon DH, Rickard JW, Kanj M, Wazni OM, Saliba WI, Varma N, et al. Incidence and predictors of right ventricular pacing-induced cardiomyopathy in patients with complete atrioventricular block and preserved left ventricular systolic function. Heart Rhythm 2016;13:2272–2278. doi: 10.1016/j.hrthm.2016.09.027

3. Cho EJ, Park SJ, Park KM, On YK, Kim JS. Paced QT interval as a risk factor for new-onset left ventricular systolic dysfunction and cardiac death after permanent pacemaker implantation. Int J Cardiol 2016;203:158–163. doi: 10.1016/j.ijcard.2015.10.128

4. Boriani G, Vitolo M, Proietti M. Cardiomyopathy associated with long-term right ventricular pacing: an intriguing clinical issue. Eur J Heart Fail 2019;21:652–654. doi: 10.1002/ejhf.1449

5. Ebert M, Jander N, Minners J, Blum T, Doering M, Bollmann A, Hindricks G, Arentz T, Kalusche D, Richter S. Long-Term Impact of Right Ventricular Pacing on Left Ventricular Systolic Function in Pacemaker Recipients With Preserved Ejection Fraction: Results From a Large Single-Center Registry. J Am Heart Assoc 2016;5. doi: 10.1161/jaha.116.003485

6. Dor O, Haim M, Barrett O, Novack V, Konstantino Y. Incidence and Clinical Outcomes of Pacing Induced Cardiomyopathy in Patients With Normal Left Ventricular Systolic Function and Atrioventricular Block. Am J Cardiol 2020;128:174–180. doi: 10.1016/j.amjcard.2020.05.017

7. Khurshid S, Epstein AE, Verdino RJ, Lin D, Goldberg LR, Marchlinski FE, Frankel DS. Incidence and predictors of right ventricular pacing-induced cardiomyopathy. Heart Rhythm 2014;11:1619–1625. doi: 10.1016/j.hrthm.2014.05.040

8. Wilkoff BL, Cook JR, Epstein AE, Greene HL, Hallstrom AP, Hsia H, Kutalek SP, Sharma A, Dual C, Investigators VVIIDT. Dual-chamber pacing or ventricular backup pacing in patients with an implantable defibrillator: the Dual Chamber and VVI Implantable Defibrillator (DAVID) Trial. JAMA 2002;288:3115–3123. doi: 10.1001/jama.288.24.3115

9. Sweeney MO, Bank AJ, Nsah E, Koullick M, Zeng QC, Hettrick D, Sheldon T, Lamas GA, Search AVE, Managed Ventricular Pacing for Promoting Atrioventricular Conduction T. Minimizing ventricular pacing to reduce atrial fibrillation in sinus-node disease. N Engl J Med 2007;357:1000–1008. doi: 10.1056/NEJMoa071880

10. Nielsen JC, Thomsen PE, Hojberg S, Moller M, Vesterlund T, Dalsgaard D, Mortensen LS, Nielsen T, Asklund M, Friis EV, et al. A comparison of single-lead atrial pacing with dual-chamber pacing in sick sinus syndrome. Eur Heart J 2011;32:686–696. doi: 10.1093/eurheartj/ehr022

11. Sagar S, Shen WK, Asirvatham SJ, Cha YM, Espinosa RE, Friedman PA, Hodge DO, Munger TM, Porter CB, Rea RF, et al. Effect of long-term right ventricular pacing in young adults with structurally normal heart. Circulation 2010;121:1698–1705. doi: 10.1161/CIRCULATIONAHA.109.866343

12. Boriani G, Tukkie R, Manolis AS, Mont L, Purerfellner H, Santini M, Inama G, Serra P, de Sousa J, Botto GL, et al. Atrial antitachycardia pacing and managed ventricular pacing in bradycardia patients with paroxysmal or persistent atrial tachyarrhythmias: the MINERVA randomized multicentre international trial. Eur Heart J 2014;35:2352–2362. doi: 10.1093/eurheartj/ehu165

13. Stockburger M, Boveda S, Moreno J, Da Costa A, Hatala R, Brachmann J, Butter C, Garcia Seara J, Rolando M, Defaye P. Long-term clinical effects of ventricular pacing reduction with a changeover mode to minimize ventricular pacing in a general pacemaker population. Eur Heart J 2015;36:151–157. doi: 10.1093/eurheartj/ehu336

14. Shurrab M, Healey JS, Haj-Yahia S, Kaoutskaia A, Boriani G, Carrizo A, Botto G, Newman D, Padeletti L, Connolly SJ, et al. Reduction in unnecessary ventricular pacing fails to affect hard clinical outcomes in patients with preserved left ventricular function: a meta-analysis. Europace 2017;19:282–288. doi: 10.1093/europace/euw221

15. Mei DA, Imberti JF, Vitolo M, Bonini N, Serafini K, Mantovani M, Tartaglia E, Birtolo C, Zuin M, Bertini M, et al. Systematic review and meta-analysis on the impact on outcomes of device algorithms for minimizing right ventricular pacing. Europace 2024;26. doi: 10.1093/europace/euae212

16. Choi EK. Cardiovascular Research Using the Korean National Health Information Database. Korean Circ J 2020;50:754–772. doi: 10.4070/kcj.2020.0171

17. Choi YJ, Choi EK, Han KD, Jung JH, Park J, Lee E, Choe W, Lee SR, Cha MJ, Lim WH, et al. Temporal trends of the prevalence and incidence of atrial fibrillation and stroke among Asian patients with hypertrophic cardiomyopathy: A nationwide population-based study. Int J Cardiol 2018;273:130–135. doi: 10.1016/j.ijcard.2018.08.038

18. Park J, Kwon S, Choi E-K, Choi Y-j, Lee E, Choe W, Lee S-R, Cha M-J, Lim W-H, Oh S. Validation of diagnostic codes of major clinical outcomes in a National Health Insurance database. International Journal of Arrhythmia 2019;20:1–7. doi:

19. Kang J, Kim HJ, Kim T, Lee H, Kim M, Lee SW, Kim MS, Koyanagi A, Smith L, Fond G, et al. Prenatal opioid exposure and subsequent risk of neuropsychiatric disorders in children: nationwide birth cohort study in South Korea. BMJ 2024;385:e077664. doi: 10.1136/bmj-2023-077664

20. Park J, Kim A, Bell ML, Kim H, Lee W. Heat and hospital admission via the emergency department for people with intellectual disability, autism, and mental disorders in South Korea: a nationwide, time-stratified, case-crossover study. Lancet Psychiatry 2024;11:359–367. doi: 10.1016/S2215-0366(24)00067-1

21. Rebora P, Salim A, Reilly M. Bshazard: a flexible tool for nonparametric smoothing of the hazard function. The R Journal 2014;6:114–122. doi:

22. Axtell AL, Bhambhani V, Moonsamy P, Healy EW, Picard MH, Sundt TM, 3rd, Wasfy JH. Surgery Does Not Improve Survival in Patients With Isolated Severe Tricuspid Regurgitation. J Am Coll Cardiol 2019;74:715–725. doi: 10.1016/j.jacc.2019.04.028

23. Snapinn SM, Jiang Q, Iglewicz B. Illustrating the impact of a time-varying covariate with an extended Kaplan-Meier estimator. The American Statistician 2005;59:301–307. doi:

24. Heinzl H, Kaider A, Zlabinger G. Assessing interactions of binary time-dependent covariates with time in cox proportional hazards regression models using cubic spline functions. Stat Med 1996;15:2589–2601. doi: 10.1002/(sici)1097-0258(19961215)15:23<2589::Aid-sim373>3.0.Co;2-o

25. Merchant FM, Hoskins MH, Musat DL, Prillinger JB, Roberts GJ, Nabutovsky Y, Mittal S. Incidence and Time Course for Developing Heart Failure With High-Burden Right Ventricular Pacing. Circ Cardiovasc Qual Outcomes 2017;10. doi: 10.1161/circoutcomes.117.003564

26. Sweeney MO, Hellkamp AS, Ellenbogen KA, Greenspon AJ, Freedman RA, Lee KL, Lamas GA, Investigators MOST. Adverse effect of ventricular pacing on heart failure and atrial fibrillation among patients with normal baseline QRS duration in a clinical trial of pacemaker therapy for sinus node dysfunction. Circulation 2003;107:2932–2937. doi: 10.1161/01.CIR.0000072769.17295.B1

27. Tayal B, Fruelund P, Sogaard P, Riahi S, Polcwiartek C, Atwater BD, Gislason G, Risum N, Torp-Pedersen C, Kober L, et al. Incidence of heart failure after pacemaker implantation: a nationwide Danish Registry-based follow-up study. Eur Heart J 2019;40:3641–3648. doi: 10.1093/eurheartj/ehz584

28. Boriani G, Savelieva I, Dan GA, Deharo JC, Ferro C, Israel CW, Lane DA, La Manna G, Morton J, Mitjans AM, et al. Chronic kidney disease in patients with cardiac rhythm disturbances or implantable electrical devices: clinical significance and implications for decision making-a position paper of the European Heart Rhythm Association endorsed by the Heart Rhythm Society and the Asia Pacific Heart Rhythm Society. Europace 2015;17:1169–1196. doi: 10.1093/europace/euv202

29. Jankowski J, Floege J, Fliser D, Böhm M, Marx N. Cardiovascular Disease in Chronic Kidney Disease: Pathophysiological Insights and Therapeutic Options. Circulation 2021;143:1157–1172. doi: 10.1161/circulationaha.120.050686

30. Liao JN, Chao TF, Tuan TC, Kong CW, Chen SA. Long-term outcome in patients receiving permanent pacemaker implantation for atrioventricular block: Comparison of VDD and DDD pacing. Medicine (Baltimore) 2016;95:e4668. doi: 10.1097/md.0000000000004668

31. Krzemień-Wolska K, Tomasik A, Nowalany-Kozielska E, Jacheć W. Prognosis of patients with implanted pacemakers in 4-year follow-up: Impact of right ventricular pacing site. Herz 2018;43:315–324. doi: 10.1007/s00059-017-4561-6

32. Riesenhuber M, Spannbauer A, Rauscha F, Schmidinger H, Boszotta A, Pezawas T, Schukro C, Gwechenberger M, Stix G, Anvari A, et al. Sex Differences and Long-Term Outcome in Patients With Pacemakers. Front Cardiovasc Med 2020;7:569060. doi: 10.3389/fcvm.2020.569060

33. Varma N, Lappe J, He J, Niebauer M, Manne M, Tchou P. Sex-Specific Response to Cardiac Resynchronization Therapy: Effect of Left Ventricular Size and QRS Duration in Left Bundle Branch Block. JACC Clin Electrophysiol 2017;3:844–853. doi: 10.1016/j.jacep.2017.02.021

34. Chen HC, Liu WH, Tseng CH, Chen YL, Lee WC, Fang YN, Chong SZ, Chen MC. Diabetes Increases Risk of Cardiovascular Events in Patients Receiving Permanent Pacemaker: A Propensity Score-Matched Cohort Study. J Diabetes Res 2022;2022:6758297. doi: 10.1155/2022/6758297

35. Kirkman MS, Briscoe VJ, Clark N, Florez H, Haas LB, Halter JB, Huang ES, Korytkowski MT, Munshi MN, Odegard PS, et al. Diabetes in older adults. Diabetes Care 2012;35:2650–2664. doi: 10.2337/dc12-1801

36. Merchant FM, Mittal S. Pacing induced cardiomyopathy. J Cardiovasc Electrophysiol 2020;31:286-292. doi: 10.1111/jce.14277

37. Tokavanich N, Prasitlumkum N, Mongkonsritragoon W, Cheungpasitporn W, Thongprayoon C, Vallabhajosyula S, Chokesuwattanaskul R. A network meta-analysis and systematic review of change in QRS duration after left bundle branch pacing, His bundle pacing, biventricular pacing, or right ventricular pacing in patients requiring permanent pacemaker. Sci Rep 2021;11:12200. doi: 10.1038/s41598-021-91610-8

38. Sze E, Samad Z, Dunning A, Campbell KB, Loring Z, Atwater BD, Chiswell K, Kisslo JA, Velazquez EJ, Daubert JP. Impaired Recovery of Left Ventricular Function in Patients With Cardiomyopathy and Left Bundle Branch Block. J Am Coll Cardiol 2018;71:306–317. doi: 10.1016/j.jacc.2017.11.020

39. Khurshid S, Obeng-Gyimah E, Supple GE, Schaller R, Lin D, Owens AT, Epstein AE, Dixit S, Marchlinski FE, Frankel DS. Reversal of Pacing-Induced Cardiomyopathy Following Cardiac Resynchronization Therapy. JACC Clin Electrophysiol 2018;4:168–177. doi: 10.1016/j.jacep.2017.10.002

40. Heidenreich PA, Bozkurt B, Aguilar D, Allen LA, Byun JJ, Colvin MM, Deswal A, Drazner MH, Dunlay SM, Evers LR, et al. 2022 AHA/ACC/HFSA Guideline for the Management of Heart Failure: A Report of the American College of Cardiology/American Heart Association Joint Committee on Clinical Practice Guidelines. Circulation 2022;145:e895–e1032. doi: 10.1161/cir.0000000000001063

41. Wang Y, Zhou R, Lu C, Chen Q, Xu T, Li D. Effects of the Angiotensin-Receptor Neprilysin Inhibitor on Cardiac Reverse Remodeling: Meta-Analysis. J Am Heart Assoc 2019;8:e012272. doi: 10.1161/jaha.119.012272

